# RELIABILITY METHODS FOR ANALYZING COVID-19 PANDEMIC SPREADING BEHAVIOR, LOCKDOWN IMPACT AND INFECTIOUSNESS

**DOI:** 10.1101/2021.04.22.21255946

**Authors:** Alicia Puls, Stefan Bracke

## Abstract

In 2021, the COVID-19 pandemic continues to challenge the globalized world. Restrictions on the public life and lockdowns of different characteristics define the life in many countries. This paper focuses on the first year of the COVID-19 pandemic (01-28-2020 to 01-15-2021). As a transfer of methods used in reliability engineering for analyzing occurrence of infection, Weibull distribution models are used to evaluate the spreading behavior of COVID-19.

Key issues of this study are the differences of spreading behavior in first and second pandemic phase and the various impacts of lockdown measures with different characteristics (hard, light). Therefore, the occurrence of infection in normed time periods with and without lockdown measures are analyzed in detail on the example of Germany representing the spreading behavior in Europe. Additional information in comparison to classical infection analyzes models like SIR model is generated by the application of Weibull distribution models with easy interpretable parameters and the dynamic development of COVID-19 is outlined.

In a further step, the occurrence of infection of COVID-19 is put into the context of other common infectious diseases in Germany like Influenza or Norovirus to evaluate the infectiousness. Differences in spreading behavior of COVID-19 in comparison to these well-known infectious diseases are underlined for different pandemic phases.

## 1. Introduction

In December 2019, the world was confronted with the outbreak of the respiratory COVID-19 (“Corona”). The first infection – confirmed case – was detected in the City Wuhan, Hubei, China. First, it was an epidemic in China, but in the first quarter of 2020 it evolved into a worldwide pandemic. The COVID-19 pandemic is caused by Coronavirus SARS-CoV-2.

In 2021, the COVID-19 pandemic continues to challenge the globalized world. Restrictions on the public life and lockdowns of different characteristics define the life in many countries. This paper focuses on the first year of the COVID-19 pandemic (01-28-2020 to 01-15-2021). The data status is March 17th 2021.

COVID-19 is an infectious disease affecting the respiratory tract. In comparison to other infectious diseases, the virus occurs worldwide, even in hot countries, and shows a low seasonal impact. Furthermore, the number of cases is much higher than the cases of other infectious diseases. In this paper, the spreading behavior of COVID-19 is compared with other infectious diseases. Moreover, the spreading behavior of the first and second COVID-19 wave is analyzed, subsequently the impact of different lockdown measures is evaluated. The focus of the quantitative analyze of the COVID-19 spreading behavior and speed is the use of Weibull distribution models.

This paper is the continuation of previous research studies, cf. Bracke et al. (2020) and Puls and Bracke (2020), where the focus is on a detailed evaluation of the characteristics of COVID-19 spreading behavior in the different pandemic phases on the example of Germany as a reference country.

## 2. Goal of Research Study

The overarching goal of the research study is the analysis of the development of infection occurrence of COVID-19 in the first year of the pandemic (01-2020 to 01-2021) with reliability methods. The detailed goals are as follows:

1. Comparison of the spreading behavior in the first and second wave,
2. Analyses of the lockdown impact with comparing different lockdown characteristics,
3. Analyses of the infectiousness of COVID-19 in different pandemic phases in relation to other already well-known infectious diseases.

These topics are discussed based on Germany as a reference country for the occurrence of infection in Europe. The reference country is selected due to data quality and access and the different lockdown characteristics. The COVID-19 spreading behaviour is compared with the most frequently occurring notifiable infectious diseases in Germany, Influenza, Norovirus and Campylobacter enteritis.

## 3. Data Base

The base of operations for the presented research study are the infection data documentation of the Johns Hopkins University (JHU). The COVID-19 dashboard by the Center for Systems Science and Engineering (CSSE) at Johns Hopkins University (2020) documents confirmed cases, recovered cases as well as death cases regarding countries and regions, starting at 01-22-2020, cf. JHU (2021). For this study, the daily confirmed cases are in focus.

As a frame for the data basis for the presented study, Table 1 shows an overview of key dates. Relevant are the dates of first infection as the beginning of the first wave, the beginning of the first, light and second lockdown and the beginning of the second wave of COVID-19 in Germany.

**Table 1.** COVID-19 development in Germany. Important phases and dates.

| Phase | Date |
| --- | --- |
| first infection | 01-28-2020 |
| first lockdown | 03-22-2020 |
| begin second wave | 09-09-2020 |
| lockdown light | 11-02-2020 |
| second lockdown | 12-16-2020 |

Note: The date for the beginning of the second wave is a defined date to get a comparable time span of 55 days from first infection to first lockdown as well as second wave to lockdown light. An alternative method would be a trend detection, e.g., with a Cox and Stuart Trend test, cf. Cox and Stuart (1955).

While the first and second wave are characterized by a more or less unhindered spread, during the lockdown hard measures were applied in part. The concrete characteristics of the different lockdowns are described in section 5.2.

For the analyses of the infectiousness of COVID-19, the comparison data of other infectious diseases is obtained from the Robert Koch Institute (RKI). The RKI is the central registration authority for notifiable diseases. Number of cases by season week per year can be accessed there for several diseases, cf. RKI (2021).

It must be considered, that there are uncertainty factors in the data base. Here a brief overview of the uncertainty with regard to data acquisition is given, for a detailed overview cf. Bracke et al. (2020). First of all, the type of measuring method has to be considered. Three aspects are important:

- Criteria for testing (test strategy, e.g., symptom-based or area-wide),
- Reporting system (reporting procedure),
- Accessibility of health department (e.g., weekend-impact).

Furthermore, apart from the lockdown measures taken, the spreading is also influenced by the dynamic occurrence of infections and by the handling of the pandemic. Some of these uncertainty factors are (without claiming to be conclusive):

- Seasonality and climatic effects, cf. Sajadi et al. (2020),
- Mutations of the virus,
- Type of treatment, cf. Gattinoni et al. (2020) and
- Vaccination progress.

These uncertainties form the framework for the comparative data, which means:

- All analyses are carried out using Germany as a reference example. Uncertainty factors like population density, different definitions of number of cases or cultural differences are avoided.
- The data is differentiated between before and after lockdown.
- Ranked data is used and the time is normalized to the date of occurrence.

In addition, the results are checked for plausibility and uncertainties during the analyses.

## 4. Method

The analyses of the spreading behavior, the lockdown impact and the infectiousness of COVID-19 in the different pandemic phases are carried out by a comparison of the Weibull distribution parameters and the Weibull probability plot. This section shows the fundamentals of the Weibull distribution model in Sec. 4.1 and the transfer to the occurrence of infection in Sec. 4.2.

### 4.1. Weibull distribution model

The two-parameter Weibull distribution model is given based on Eq. (1), cf. Weibull (1951).

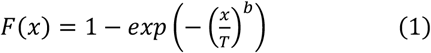

The parameters, besides the term life span variable x, are scale parameter T (in lifetime analysis: characteristic life span) and shape parameter b. By variating parameter b, different failure rates can be described, therefore the Weibull model can be flexibly used for different applications, cf. Rinne (2008). The shape parameter b gives hints regarding the character of the failure period: early failure period, random failure or operation time related failure behavior. The Weibull parameters are estimated by using the Maximum Likelihood Estimator (MLE), cf. Fisher (1912).

### 4.2. Transfer to occurrence of infection

The Weibull distribution model is frequently used within reliability engineering and risk analytics, cf. Birolini (2017). The exponential spreading of case numbers with a saturation limit enables the application of this model to the analyses of the occurrence of infection. In addition to classical methods of virology such as the SIR model (cf. Kermack and McKendrik (1927)), e.g., applied for COVID-19 in D’Arienzo and Coniglio (2020) in combination with basic reproduction number, the Weibull distribution model offers the possibility, to gain knowledge with regard to the infection development respectively the infection gradient. One advantage of the Weibull distribution model in comparison to other distribution models, e.g., the exponential distribution, are the easy interpretable model parameters. In occurrence of infection, the shape parameter b as gradient of the model is interpreted as spreading speed (transfer thinking of reliability engineering: shape parameter b describes the occurrence of damage cases within a product fleet). The scale parameter T gives another hint of the spreading speed considering the first infection case. Representing the x value of the probability 0.63, T indicates while comparing different models (e.g. different pandemic phases) how fast the infection cases are progressing in relation to the total days (Puls and Bracke 2020). In Figure 1 the daily confirmed COVID-19 cases in Germany of the first 100 days of the pandemic are shown as there occur in the JHU database.

**Fig. 1:**
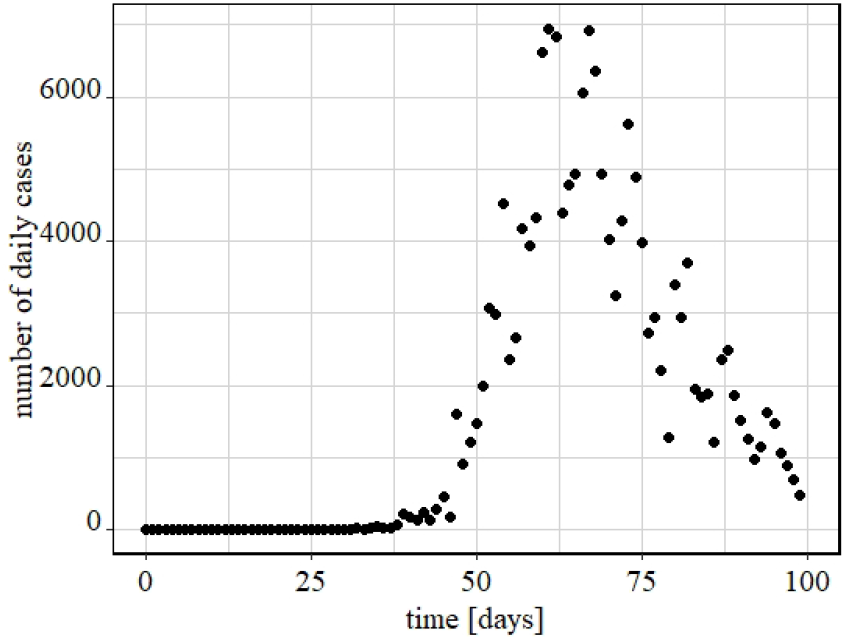
Daily confirmed cases of COVID-19 in Germany in the first 100 days of the pandemic.

A time span of about 40 days where hardly any infections occur can be identified. After this steady phase, there is an exponential increase of case numbers until day 65. Then the number of cases is decreasing.

For the application of the Weibull distribution model the data of infection must be ranked by days. The Weibull distribution model requires occurrence times as an input variable; for the infection data, the occurrence time is the reported infection point of time. To avoid a mixture distribution, every pandemic phase like first wave (increasing case numbers) or first lockdown (decreasing case numbers) are ranked separately. For comparability, the start day of every pandemic phase is set to “day one” in the data set.

## 5. Data Analytics

### Spreading behaviour, Lockdown impact, Infectiousness

This section focused on COVID-19 data analytics. The spreading behavior of first and second wave is compared, cf. Sec. 5.1. Furthermore, the impact of the different lockdowns on the occurrence of infection is evaluated in Sec. 5.2. Subsequently, the infectiousness of COVID-19 is put into relation with other infectious diseases in first and second pandemic phase, cf. Sec. 5.3. Therefore, Weibull distribution models with probability plots and model parameters with confidence belts are analyzed.

### 5.1. Spreading behavior

In this section, the spreading behavior of COVID-19 in Germany without lockdown impact is analyzed for the first and second pandemic phase. The first wave of COVID-19 started at January 28th, 2020; the point of time of the first lockdown was on March 22nd, 2020, cf. Tab. 1. This results in a time span of 55 days more or less undisturbed spreading (without any breaking effect or measure).

To enable a meaningful comparison of the spreading behavior of first and second pandemic phase, the time span for the second wave is set to 55 days as well. Therefore, as a start point of the second wave the September 9th, 2020 is chosen as the date 55 days before the start of the lockdown light measure on November 2nd, 2020, cf. Tab. 1. The fit of the Weibull distribution models as described in Sec. 4.2, results in the Weibull plot shown in Figure 2 and the related Weibull parameters in Table 2.

**Table 2.** Weibull model parameters first and second wave (cumulative confirmed cases). Confidence level γ = 0.95.

| phase | cases | $T$ [d] | shape $b$, confidence belt |
| --- | --- | --- | --- |
| first wave | 24,913 | 46 - | $17.83 \leq 18.00 \leq 18.18$ |
| second wave | 314,641 | 46 | $3.544 \leq 3.554 \leq 3.564$ |

**Fig. 2:**
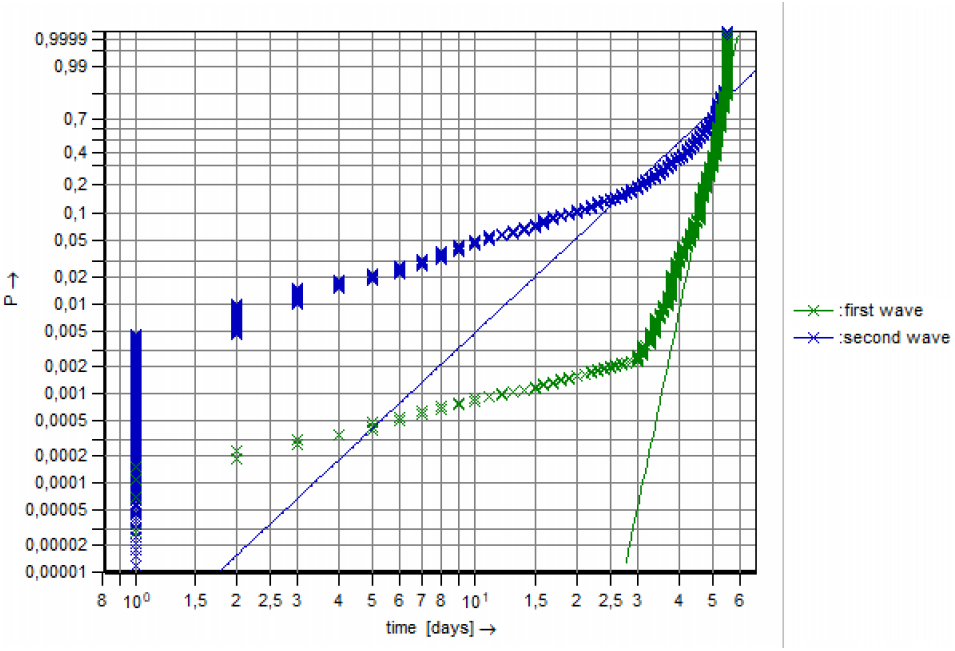
Weibull distribution models COVID-19, comparison first and second wave, confirmed cases, time span 55 days.

First of all, in Figure 1 the difference of the spreading behavior of COVID-19 in Germany between first and second wave gets clear. The curve of the first wave, plotted in green color, has a greater slope. The gradient, expressed by the shape parameter is larger. Therefore, the first wave is characterized by a higher spreading speed in comparison to the second wave.

These findings are confirmed by the comparison of the shape parameters in Table 2. The shape parameter of the first wave is about five time higher than the shape parameter of the second wave. When comparing these shape parameters with results in reliability, the shape parameter of the first wave stands out as well. Typical wear out mechanism in mechanical engineering are located between 1.5 ≤ b ≤ 3, brittle failures have typically shapes of b ~ 8. As a transfer on occurrence of infection: The first wave of COVID-19 shows a strong gradient respectively spreading speed.

The scale parameter for the first and second wave is the same. A difference is present for the number of cases, there the case number in the second wave is greater. This represents the progressed occurrence of infection in the second wave. The advantage of the use of Weibull distribution models as comparison is the normalization, so the analysis of spreading speed is possible despite the different number of cases in the two periods under consideration.

### 5.2. Lockdown impact

In Germany, with status March 2021 three lockdown measures were executed. The first lockdown took place in the first phase of the pandemic, from March 22nd, 2020 to May 3rd, 2020. In the second wave there was at first a “lockdown light” from November 2nd, 2020 to December 14th, 2020, which was extended to the second lockdown from December 16th, 2020 to first easing of measures on March 8th, 2021. The lockdown measures have different characteristics, cf. Table 3. As columns the different lockdown measures (first, light, second lockdown) are described, as rows the concrete measures. Different characteristics are highlighted.

**Table 3.** Lockdown (LD) characteristics in Germany, differences are highlighted.

| measure | first LD | LD light | second LD |
| --- | --- | --- | --- |
| hygiene | yes | yes | yes |
| none mayor events | yes | yes | yes |
| shutdown of educational system | yes | **no** | yes |
| border controls | yes | **no** | yes |
| shutdown of retail | yes | **no** | yes |
| distance regulations | yes | yes | yes |
| contact restrictions | **no** | yes | yes |
| masks | **no** | yes | yes |

Hygiene measures and the prohibition of mayor events were applied in every lockdown. While comparing the first and second lockdown, measures like contact restrictions and masks appear. During the first lockdown, these measures were not used. Therefore, contact restrictions and masks were applied in the lockdown light as well. The lockdown light had a soft impact on social life, because there was no shutdown of educational systems, border controls nor the shutdown of retail.

For the comparison of the spreading behavior with regard to different lockdown measures, the time span is normalized on about 42 days, the duration of the first and the lockdown light. Therefore, the period under review of the second lockdown is shortened from December 16th, 2020 to January 27th, 2021. Weibull distribution models are fitted, as a result these are shown as probability plots in combination with the spreading behavior of the first and second wave in Figure 3. The corresponding Weibull parameters are documented in Table 4.

**Table 4.** Weibull model parameters first and second wave with lockdown impact (cumulative confirmed cases). Confidence level γ = 0.95.

| phase | cases | T [d] | shape b, confidence belt |
| --- | --- | --- | --- |
| first wave | 24,913 | 46 - | $17.83 \leq 18.00 \leq 18.18$ |
| first lockdown | 140,791 | 17 | $1.501 \leq 1.507 \leq 1.513$ |
| second wave | 314,641 | 46 | $3.544 \leq 3.554 \leq 3.564$ |
| lockdown light | 806,464 | 24 | $1.681 \leq 1.683 \leq 1.686$ |
| second lockdown | 772,957 | 21 | $1.493 \leq 1.496 \leq 1.498$ |

**Fig. 3:**
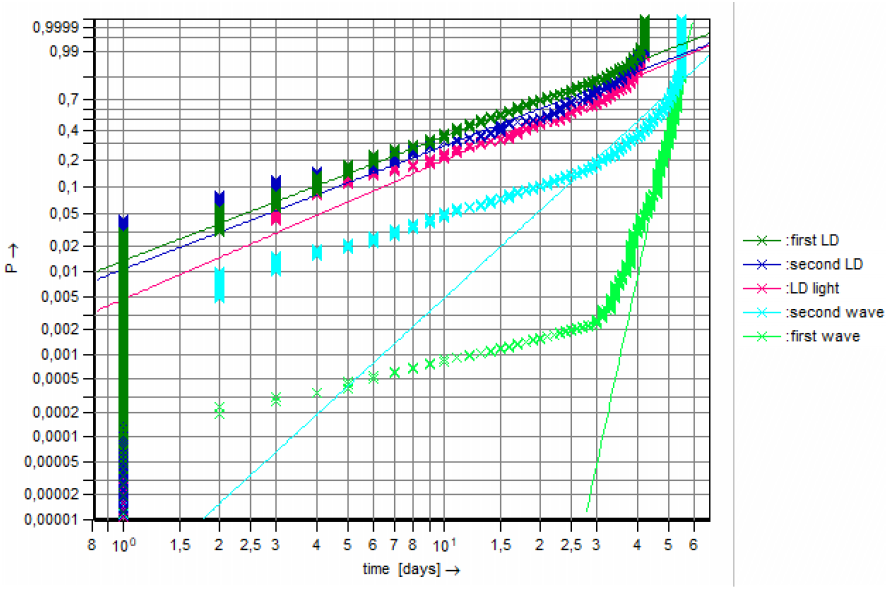
Weibull distribution models COVID-19 first and second wave, confirmed cases, lockdown (LD) impact, time span 42 resp. 56 days.

A clear difference can be noticed between the spreading behavior with lockdown (first, light, second lockdown) and without lockdown (first, second wave). The gradient of the Weibull models is significantly reduced, caused by the lockdown measures. The gradient of the lockdown light is slightly greater than the gradient of the first and second lockdown. Between the first and second lockdown, no significant difference can be noticed by comparing the curves in Figure 3 respectively the gradient (shape b) in Table 4.

While comparing the shape parameters in Table 4, the significant impact of all lockdown measures gets clear. The first lockdown reduced the shape parameter respectively the spreading speed with almost factor 12. The lockdown light and the second lockdown reduced the spreading speed of the second wave with factor ~2. It has to be noticed, that the spreading speed of the second wave was much lower than the spreading speed in the first wave. Therefore, it can be assumed, that the effect of the lockdown measures in the second pandemic phase, was not as relevant as in the first phase.

The comparison of the COVID-19 spreading speed within the three lockdown phases results in the following ranking; cf. Table 4: During the second lockdown, the spreading speed was on the lowest level. The spreading speed within the first lockdown was higher in comparison to the second lockdown phase. Furthermore, the spreading speed within the lockdown light phase was on the highest level. The ranking is significant, based on the consideration of the confidence belt (cf. Table 4). This observation fits with the different characteristics of the three lockdowns measures, cf. Table 3. During the second lockdown phase, the most measures were valid. The first lockdown phase contains the second most measures and in the lockdown light not all measures were applied. It is stated that a lockdown with a shutdown of educational system and retail in combination with border controls, distance and contact restrictions and masks correlates with the lowest observed COVID-19 spreading speed.

The differences of the scale parameter T in Table 4 are related to the different time spans (first and second wave: 55 days and lockdown measures: 42 days). They do not affect the results of the spreading behavior. Also, the different number of cases do not influence the spreading speed due to the normalization of the Weibull distribution model, cf. Sec. 5.1. The different pandemic phases can be characterized by the shown parameters, especially the spreading speed (gradient respectively shape parameter b). Besides it can be observed, that the number of cases within the lockdown light phase are on a higher level in comparison to the second wave phase. Hence, the lockdown light measure had the effect of the prevention of a (further) exponential increase regarding the number of cases.

### 5.3. Infectiousness

For an assessment of the COVID-19 pandemic, the spreading behavior is compared with other common notifiable diseases in Germany: Influenza, Norovirus and Campylobacter Enteritis. First, the seasonal beginnings of these diseases are compared with the beginning (first wave) of COVID-19, cf. Sec. 5.3.1. In a second step, the peak (worst case of the spreading behavior) is compared with the second wave of COVID-19 in Section 5.3.2.

Influenza is a seasonal (Winter half-year) respiratory infectious disease with symptoms like fever, cough, sore throat or muscular pains. Infection occurs via droplets from person to person. The seasonal number of cases in Germany varies between 3,000 and 270,000 cases. (RKI 2019) Norovirus is a seasonal (winter months) gastro-intestinal disease with vomiting and diarrhea. The infection is fecal-oral or by droplets from person to person. The number of cases in Germany range between 60,000 and 100,000 cases per season. (RKI 2019)

Campylobacter Enteritis (CE) is also a gastrointestinal disease. It occurs seasonally in warm season and shows symptoms like fever, diarrhea and stomach ache. Unlike the Norovirus, this infectious disease is spread through food. The transmission from person to person is rather rare. The seasonal number of cases in Germany range between 50,000 and 70,000 cases. (RKI 2019)

#### 5.3.1. Infectiousness in first pandemic phase

The comparison data set of the other infectious diseases (RKI 2021) is given on a weekly base by season. For a meaningful comparison, a five-year-mean for each season week is estimated in order to avoid outliers. Therefore, the seasons from 2014/15 to 2018/19 are analysed. The season 2019/20 is not considered for the comparison because of the interdependency to the concurrently valid COVID-19 protective measures.

To get a sound basis for comparison with the first wave of COVID-19 (55 days), the first eight weeks (56 days) of each disease are analyzed. According to the weekly data base, a transformation one week to seven days is made resulting in eight nodes for the Weibull distribution model fits regarding to the other infectious diseases. Based on the ranked data (cf. Sec. 4.2) a Weibull plot and the corresponding Weibull parameters are estimated (cf. Figure 4 and Table 5). Figure 4 shows the spreading behavior regarding the COVID-19 first wave, COVID-19 first lockdown, and the infectious diseases Influenza, Norovirus, Campylobacter Enteritis.

**Table 5.** Weibull model parameters COVID-19 and other infectious diseases in first wave (cumulative confirmed cases). Confidence level γ = 0.95.

| phase/disease | cases | T [d] | shape b<br>[confidence belt] |
| --- | --- | --- | --- |
| first wave<br>COVID-19 | 24,913 | 46 - | $17.83 \leq 18.00 \leq 18.18$ |
| first lockdown<br>COVID-19 | 140,791 | 17 | $1.501 \leq 1.507 \leq 1.513$ |
| CE | 10,660 | 35 | $2.034 \leq 2.065 \leq 2.097$ |
| Influenza | 277 | 45 | $2.676 \leq 2.950 \leq 3.237$ |
| Norovirus | 11,124 | 42 | $2.596 \leq 2.635 \leq 2.675$ |

**Fig. 4:**
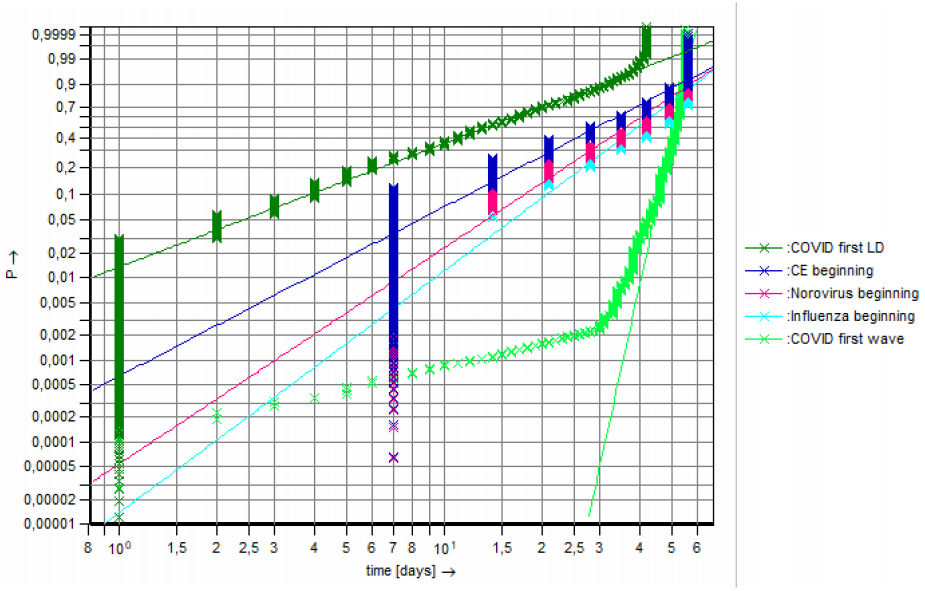
Weibull distribution models COVID-19 and other infectious diseases (5-year-mean 2014/15 to 2018/19) in first wave/beginning, confirmed cases, time span 56 days.

It gets clear that the unhindered spreading speed of COVID-19 is on a much higher level in comparison to the infectious diseases Influenza, Norovirus, Campylobacter Enteritis (CE). The curve of COVID-19 first wave shows a significant different characteristic than the curves of the other infectious diseases. While comparing the shape parameters (cf. Table 5), a factor of ~9 (CE) or ~6 (Influenza) occurs in comparison of the shape parameter (spreading speed) of COVID-19. The spreading speed of COVID-19 at the beginning of the pandemic is essentially higher than the spreading speed of “normal” infectious diseases like Influenza or Norovirus. Dismissing the COVID-19 spreading as the spreading of e.g., a “normal Influenza” is therefore wrong and hazardous.

With the lockdown impact in the first lockdown in March 2020, the spreading speed of COVID-19 is significantly reduced under the level of the unhindered speed in first wave as well under the level of the other infectious diseases. The curve of COVID-19 first lockdown (LD) in Figure 4 has a lower gradient than the other curves. The comparison of shape parameters also shows a significant lower spreading speed within the first lockdown phase. With the strict lockdown measures like shutdown of educational systems, shutdown of retail, border controls and hygiene measures, the spreading speed of COVID-19 can be reduced to a level of the spreading speed of the infectious diseases Influenza, Norovirus, Campylobacter Enteritis (CE). In this case, the spreading speed of COVID-19 is slightly lower than the spreading speed of other infectious diseases like Influenza without measures. This outlines again the effectiveness of the lockdown measures to control the COVID-19 pandemic in the first wave.

#### 5.3.2. Infectiousness in second pandemic phase

To evaluate the infectiousness of COVID-19 in the second wave, a comparison with the peak of Influenza spreading behavior is made. This peak is identified by the maximum of the weekly cases in the five-year-mean season of Influenza. The review period are the eight weeks (56 days) before this peak to generate a “worst case spreading behavior” as a base of comparison with the second wave of COVID-19 (55 days) and the light and second lockdown of COVID-19 in the end of 2020, cf. Table 1. The resulting Weibull plot is shown in Figure 5 and the corresponding Weibull parameters are documented in Table 6.

**Table 6.**
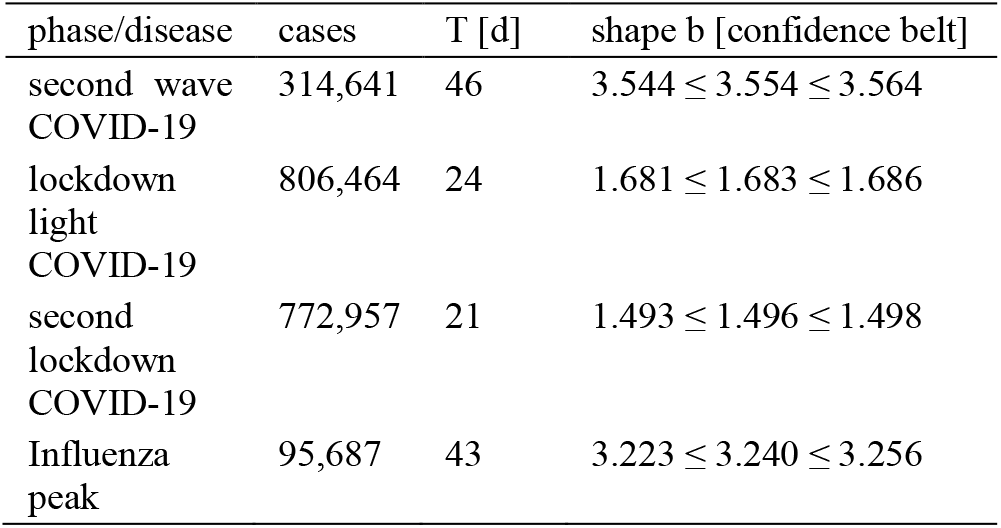
Weibull model parameters COVID-19 and Influenza in second wave (cumulative confirmed cases). Confidence level γ = 0.95.

| phase/disease | cases | T [d] | shape b [confidence belt] |
| --- | --- | --- | --- |
| second wave<br>COVID-19 | 314,641 | 46 | $3.544 \leq 3.554 \leq 3.564$ |
| lockdown<br>light<br>COVID-19 | 806,464 | 24 | $1.681 \leq 1.683 \leq 1.686$ |
| second<br>lockdown<br>COVID-19 | 772,957 | 21 | $1.493 \leq 1.496 \leq 1.498$ |
| Influenza<br>peak | 95,687 | 43 | $3.223 \leq 3.240 \leq 3.256$ |

**Fig. 5:**
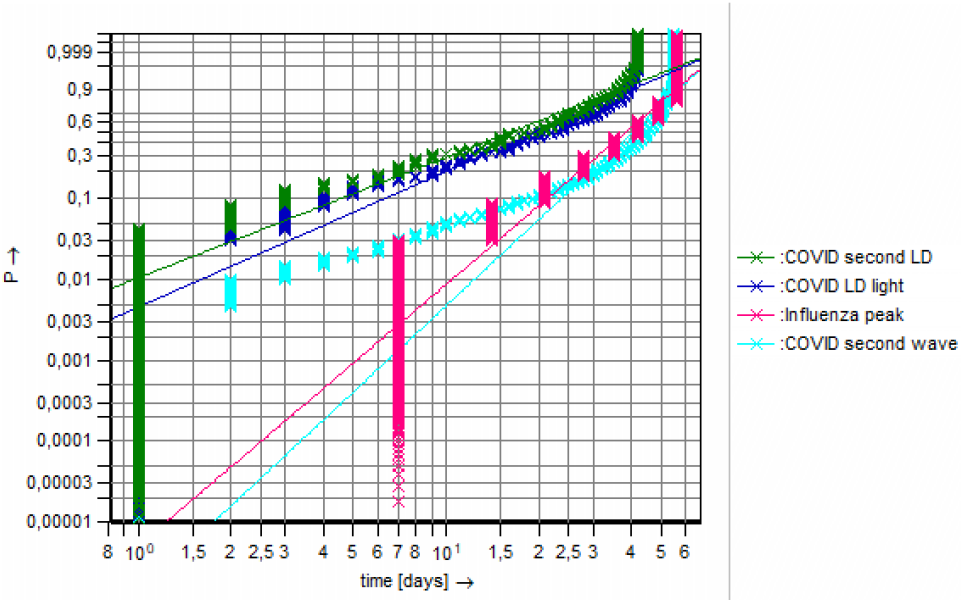
Weibull distribution models COVID-19 and Influenza (5-year-mean 2014/15 to 2018/19) in second wave/peak, confirmed cases, time span 56 days.

In the second COVID-19 pandemic phase, the spreading speed of is only slightly higher than the spreading speed of other infectious diseases, here represented by Influenza. It has to be considered, that the comparison base of Influenza is a “worst case scenario” without control measures while the COVID-19 spreading in the second wave was accompanied with measures like distance regulations and masks. However, the shape parameter representing the spreading speed of the COVID-19 second wave is significantly higher than the parameter of the Influenza peak phase. It is remarkable, that the spreading speed of the “worst case scenario” of Influenza is still the factor ~6 times lower than the COVID-19 spreading speed of in first wave, cf. Table 5 and 6.

Both the lockdown light and the second lockdown in the second pandemic phase reduce the spreading speed of COVID-19 under the level of the other infectious diseases, here represented by Influenza. Figure 5 shows the impact of lockdown measures regarding the slowdown of the COVID-19 pandemic.

What has to be noticed in the evaluation of the infectiousness of COVID-19 in the first and second pandemic phase are the extreme higher number of confirmed cases both with and without lockdown measures in comparison to other infectious diseases, cf. Table 5 and The Weibull distribution model with the shape parameter normalizes the number of cases, therefore only the spreading speed as the gradient as the shape parameter is evaluated. With factor ~14 in the first pandemic phase and factor ~8 in the second pandemic phase of number of cases in the same period of time the infectiousness of COVID-19 gets clear.

## 6. Summary

In this paper, the method transfer from reliability engineering to epidemiology is the base for data analytics. Weibull distribution models were used for analyses of occurrence of infection and spreading behavior of COVID-19. The central aspect was the interpretation and analysing of the shape parameter b (gradient) as spreading speed. Base of operations regarding COVID-19 pandemic data are the data bases of JHU and RKI, focusing on Germany as reference country.

In the first wave in the beginning of 2020, COVID-19 showed a strong increase of number of cases. The spreading speed, represented by the shape parameter of the Weibull distribution model, is extraordinary in comparison to typical technical damage cases in the product use phase within reliability analytics. The analyze of the spreading behavior in the second wave resulted in a significantly lower spreading speed, in contrast the number of cases in the same time span were 13 times higher than in the first wave.

All lockdown measures had a significant impact on the spreading behavior: the spreading speed slows down significantly. The highest reduction was found in the comparison of the spreading speed in the first wave phase versus in the first lockdown phase (factor around 12). The most efficient lockdown measure was the second lockdown in the end of 2020, followed by the first lockdown and the least efficient lockdown measure was the lockdown light. Lockdown measures like shutdown of educational system and retail in combination with border controls, distance and contact restrictions as well as masks reduce the spreading speed very strong.

For the evaluation of infectiousness, the COVID-19 spreading behavior was set into relation with other common infectious diseases in Germany (Influenza, Norovirus and Campylobacter Enteritis). In a first step, the COVID-19 first wave was compared with the season beginnings of the other diseases. The COVID-19 spreading behavior is not comparable with the spreading of “normal” infectious diseases like Influenza: COVID-19 had a much higher (factor around 6 to 9) spreading speed (Weibull shape parameter), only with lockdown measures the spreading behavior gets on a comparable level.

In a second step, the COVID-19 second wave was set into relation with the peak of the Influenza season as worst-case scenario of an infectious disease in Germany. COVID-19 still had a slightly faster spreading speed than Influenza (factor around 1.1), though there were measures like masks and distance regulations valid in this phase of the pandemic.

Notable is the fact, that the worst-case Influenza phase shows still a much lower spreading speed (factor around 5.6) than COVID-19 in the first wave. The lockdown light and the second lockdown both reduced the spreading speed significant as well as in comparison with the Influenza peak phase. Furthermore, the huge difference between the number of cases could be noted: There are much more (factor around 8.4) COVID-19 cases in the eight weeks data base of the second pandemic phase than Influenza cases in the peak of a five-year-mean phase. The Weibull distribution model normalizes the input variable, so this effect cannot be noticed by the shape parameter, the gradient of the model. With the knowledge of the case numbers, it can be stated, that also in the second COVID-19 pandemic phase the spreading behavior differs from the spreading behavior of other infectious diseases like Influenza.

With the use of Weibull distribution models for the analyzing of the COVID-19 pandemic additional information were generated. COVID-19 shows an extraordinary spreading behavior, especially in the first wave phase. The lockdown measures take in Germany significantly reduced the spreading speed. The spreading behavior of the COVID-19 pandemic (without any measures) is on a much higher level in comparison to the infectious diseases Influenza, Norovirus, Campylobacter Enteritis.

## Data Availability

Base of operations is the COVID-19 data base from Johns Hopkins University (JHU)

https://gisanddata.maps.arcgis.com/apps/opsdashboard/index.html#/bda7594740fd40299423467b48e9ecf6

## Notes

### Competing Interest Statement

The authors have declared no competing interest.

### Funding Statement

no funding

### Author Declarations

University of Wuppertal, Germany

